# Ventricular Dyssynchrony Late after the Fontan Operation is Associated with Decreased Survival and the Presence of Arrhythmias

**DOI:** 10.1101/2023.02.27.23286545

**Authors:** Addison Gearhart, Sunakshi Bassi, Rahul H. Rathod, Rebecca S. Beroukhim, Stuart Lipsitz, Maxwell P. Gold, David M. Harrild, Audrey Dionne, Sunil J. Ghelani

**Affiliations:** Department of Cardiology, Boston Children’s Hospital, Boston, MA, USA; Department of Pediatrics, Harvard Medical School, Boston, MA, USA; Division of General Internal Medicine, Brigham and Women’s Hospital, Boston, MA; Massachusetts Institute of Technology, Boston, MA

**Keywords:** congenital heart disease, dyssynchrony, Fontan, cardiac magnetic resonance imaging

## Abstract

**Background:** Ventricular dyssynchrony and its relationship to clinical outcomes is not well characterized in patients following Fontan palliation.

**Methods:** Single-center retrospective analysis of cardiac magnetic resonance (CMR) imaging of patients with a Fontan circulation and age-matched healthy comparison cohort as controls. Feature tracking was performed on all slices of a ventricular short-axis cine stack. Circumferential and radial strain, strain rate, and displacement were measured; and multiple dyssynchrony metrics were calculated based on timing of these measurements (including standard deviation of time-to-peak, maximum opposing wall delay, and maximum base-to-apex delay). Primary endpoint was a composite measure including time to death or heart transplant listing (D/HTx); secondary outcomes were the presence of atrial or ventricular arrhythmias.

**Results:** A total of 503 cases (15y; IQR 10, 21) and 42 controls (16y; IQR 11, 20) were analyzed. Compared to controls, Fontan patients had increased dyssynchrony metrics, longer QRS duration, larger ventricular volumes, and worse systolic function. Dyssynchrony metrics were higher in patients with right ventricular (RV) or mixed morphology compared to those with LV morphology. At median follow-up of 4.3 years, 11% had D/HTx, 7% ventricular arrhythmia, and 38% atrial arrhythmia. Multiple risk factors for D/HTx were identified, including RV morphology, ventricular dilation, dysfunction, QRS prolongation, and dyssynchrony. Ventricular dilation and RV morphology were independently associated with D/HTx; ventricular dilation and global circumferential strain were independently associated with ventricular and atrial arrhythmias.

**Conclusions:** Mechanical dyssynchrony is highly prevalent in functional single ventricles palliated to the Fontan circulation and is more pronounced in hearts with RV or mixed ventricular morphology compared to those with LV morphology. Dyssynchrony is associated with death or need for heart transplantation and cardiac arrhythmias. These data add to the growing understanding regarding factors that can be used to risk-stratify patients with the Fontan circulation.

## Introduction

Despite significant improvements in life expectancy after the Fontan operation, patients with functional single ventricles (FSV) often experience progressive heart failure and arrhythmias which may progress to death or heart transplantation (D/HTx).^1–3^ The prevalence of supraventricular and ventricular tachycardia in this population has been reported at 9% and 4%, respectively over an average follow-up of 9 years in contemporary cohorts.^4,5^ The estimated 20-year survival for patients palliated to a Fontan circulation is 61−85% with a risk of late mortality of approximately 2% per year.^6^ Sudden death is among the most common cause of mortality in this population,^7^ raising suspicion that fatal arrhythmia may be responsible,^8^ particularly since events are often not directly observed.

Synchrony plays an important role in efficient ventricular pump function and cardiac output. In a normal 2-ventricle circulation, the fast activation of electrical conduction through the heart is reflected by a narrow QRS on electrocardiogram (ECG) and a synchronous pattern of contraction. Left ventricular dyssynchrony is defined by temporal differences in activation (electrical dyssynchrony) and contraction (mechanical dyssynchrony) of various myocardial segments of the left ventricle. In adult and pediatric patient populations with biventricular circulations, ventricular dyssynchrony has been associated with life-threatening arrhythmias,^9^ heart failure,^10,11^ and mortality.^12–14^ Feature tracking (FT) to derive strain parameters has been proposed as a sensitive tool to evaluate global and regional myocardial deformation and mechanical ventricular dyssynchrony in patients with FSVs.^15^ Prior echocardiography-based studies in children with FSVs have demonstrated high rates of ventricular dyssynchrony, particularly for patients with hypoplastic left heart syndrome.^16–18^ In patients with FSVs, mechanical dyssynchrony has been associated with reduced ejection fraction (EF), longer QRS duration, and composite outcomes such as unplanned hospitalizations and/or death.^19^ However, prior studies have not provided a detailed characterization of patient-related factors that are associated with dyssynchrony (e.g., underlying ventricular morphology), and their relative contribution to patient outcomes.

The present study uses cardiac magnetic resonance (CMR) to characterize mechanical dyssynchrony in FSVs using FT. It describes novel dyssynchrony metrics and analyzes their relationship with ventricular morphology, volumetric and function data, and QRS duration. In addition, the study evaluates the association between dyssynchrony and clinical outcomes including death, heart transplantation, and arrhythmias.

## Methods

This was a single-center, retrospective cohort study. The Institutional Review Board approved the study and waived the need for informed consent.

### Study Population

All patients with a Fontan circulation who had at least one available CMR after 7/1/05 were screened for eligibility. Ventricular morphology was designated as either LV or RV, as appropriate, if the non-dominant ventricle was ≤20% the combined end diastolic volume (EDV) and mixed type if the non-dominant ventricle was >20% of the combined EDV. When multiple studies were available per patient, the earliest CMR was analyzed. Patients with inadequate image quality for FT analysis were excluded. A comparison group was identified as individuals referred for CMR for suspected cardiomyopathy or congenital heart disease (CHD) but whose studies were subsequently interpreted as normal and at the time of study acquisition or interim follow-up did not have known systemic or genetic disease. The comparison group was age-matched ∼1:12 due to the limited availability of normal studies. Demographic and clinical data were extracted from electronic medical records.

### CMR examinations

The Fontan imaging was conducted according to standard practice at our center, as has been previously described.^20^ Briefly, studies were performed on a 1.5T scanner (Achieva, Philips Healthcare, Best, the Netherlands) using surface coils appropriate for patient size. A ventricular short-axis balanced steady-state free precession (bSSFP) cine stack with breath-holding and ECG-gating was used for volumetric and FT analysis. Typical slice thickness was 8–10 mm. Typical spatial resolution was 1.7-2 × 1.7-2 mm and temporal resolution was 30-40 ms with 30 reconstructed phases per cardiac cycle. Ventricular volumes and blood flow were measured using commercially available software (cvi42, Circle Cardiovascular Imaging Inc., Calgary, Alberta, Canada; and QMass, Medis Medical Imaging Systems, Leiden, the Netherlands). The following conventional measurements were recorded: indexed end-diastolic volume (EDV_*i*_), indexed end-systolic volume (ESV_*i*_), indexed stroke volume (SV_*i*_), EF, indexed ventricular mass (Mass_i_), and ascending aortic flow as a measure of cardiac output. When two ventricles contributed to the systemic circulation (mixed type ventricles), their mass and volumes were combined. For the comparison cohort, only the LV measurements were considered.

### Feature tracking and dyssynchrony indices

Feature tracking analysis was performed on the short-axis cine stack of images as previously described.^21^ Briefly, the endocardial and epicardial borders were manually traced at end-diastole for all slices from the apex to base of the dominant ventricle in the case of a single LV or single RV. For mixed-type ventricles, borders were traced around both ventricles (excluding the ventricular septum). Contours were manually adjusted in end-diastole to ensure optimal tracking. A minimum of 4 slices was required. The apical slice was defined as the most apical slice with blood pool through the entire cardiac cycle. The basal slice was defined as the most basal slice with a full rim of myocardium through the entire cardiac cycle. Studies with artifact at the base of the heart that precluded accurate volumetric analysis were included if the most basal slice with circumferential myocardium was free of artifact and suitable for FT. For the comparison cohort, the same analysis was performed on the LV.

From this, the following six deformation measurements were obtained: circumferential strain (CS), circumferential strain rate (CSR), circumferential displacement (CD), radial strain (RS), radial strain rate (RSR), radial displacement (RD). These deformation measurements were used to calculate quantify dyssynchrony using 4 methods: 1) standard deviation of time-to-peak (SDTTP) for circumferential and radial deformation measurements for all segments, 2) maximum opposing wall delay (MOWD) as the maximum difference in the average time-to-peak for 3 opposing wall pairs, and 3) base-to-apex delay (BAD) as the time difference between peak deformation (average of 6 segments) of the most basal slice and the most apical slice (BAD1) as well as 4) the difference between the peak deformation for the most basal 2 slices and most apical 2 slices (BAD2). As such, 24 dyssynchrony indices were derived from the FT data (Figure 1/Central Illustration). Global deformation measures were recorded as global circumferential strain (GCS), global circumferential strain rate (GCSR), global radial strain (GRS), and global radial strain rate (GRSR). QRS duration was captured as a marker of electric dyssynchrony from the ECG closest to the CMR from all available ECGs, with no intervening surgical or catheter-based procedure having been performed. Heart rate was recoded from the CMR report and maximal heart rate (MHR) was calculated by using the Tanaka equation (MHR = 208 - .7*[age at CMR]). Heart rate was standardized as percent MHR (%MHR) by dividing the heart rate by MHR.^22^

**Figure 1:**
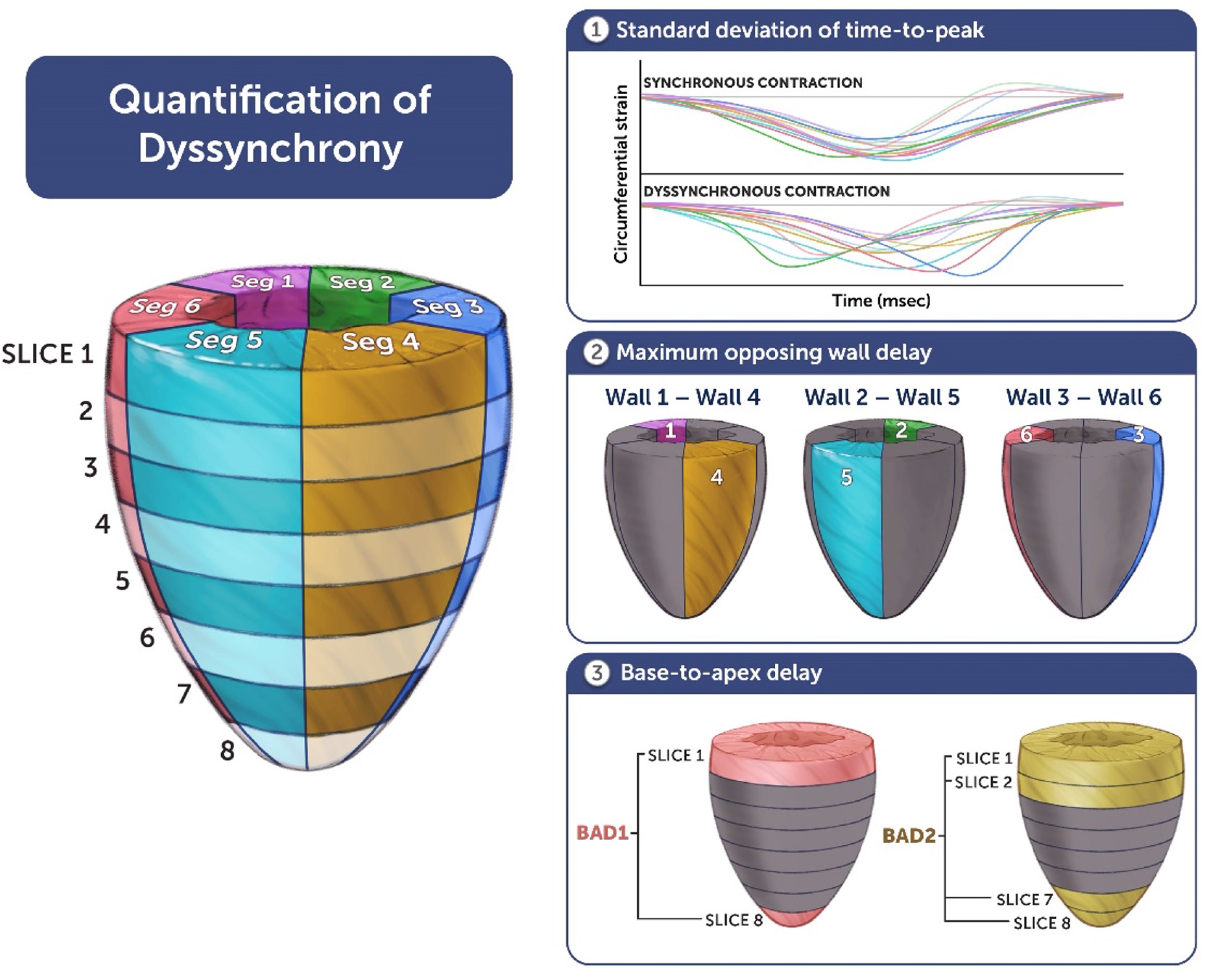
**Title:** Indices of mechanical dyssynchrony **Caption:** EDV_*i*_, indexed ventricular end-diastolic volume; EF, ejection fraction, SDTTP-CS, standard deviation time to peak circumferential strain; LV, left ventricle; RV, right ventricle. *Indicates p-value<0.05.

### Outcomes

The primary outcome was a time-to-event composite outcome of all-cause mortality or heart transplant listing (D/HTx). For time-to-event analysis, follow-up was measured from the date of CMR to the composite outcomes (earliest event in case of multiple) or last known documented follow-up in the medical record. If the first occurrence of transplant listing was prior to the CMR, it was excluded as an outcome. The secondary outcome was ventricular arrhythmias (sustained ventricular tachycardia or ventricular fibrillation), or atrial arrhythmias (atrial flutter, atrial fibrillation, or supraventricular tachycardia). Time-to-event analysis was not performed for the secondary outcome as date of first arrhythmia occurrence was not reliably well-documented.

### Statistical analysis

Data are presented as a median with interquartile ranges (IQR) as appropriate for continuous variables and as a frequency (percentage) for categorical variables. Continuous variables were compared between groups using a Mann-Whitney U test while proportions were compared using a Fisher’s exact test. Correlation between continuous variables was quantified using the Spearman’s Rho (ρ) and Pearson’s correlation coefficient (r) when the data were normally distributed. Using a set of 11 *a priori* chosen predictors, bivariate and multivariable Cox regression analyses were performed to assess relationships between predictors and time to composite outcome. Missing data for the predictors in the Cox regression model were addressed with multiple imputation.^23,24^ Based on the number of the composite outcome events, a priori, a threshold of at most 5 predictors to be included in multivariable Cox model (i.e. maximum of 1 predictor for 10 outcomes) was set. Predictors chosen for the final Cox regression model were based on a forward selection Akaike Information Criterion (AIC). The secondary outcomes of having at least one ventricular arrhythmia or atrial arrhythmia during their follow-up period are binary outcomes in the form of 0 if no arrhythmia during the follow-up period and 1 if at least 1 arrhythmia during the follow-up period (note, our arrhythmia data was collected in this dichotomized form). The binary outcomes can be considered dichotomized Poisson count outcomes of the number of events during a follow-up period, and our interest is in estimating the rates of arrhythmias per patient-days follow-up. A dichotomized Poisson variable follows a binary regression model with complementary log-log link function and the log of patient-days as an offset; the regression parameters from this binary regression model can be interpreted as log rate ratios.^25–27^ Thus, to model the rates of arrhythmias (events per patient-days follow-up), bivariate and multivariate complementary log-log binary regression models were fit to assess relationships between predictors and these arrhythmia outcomes. The maximum number of predictors was determined a priori by taking the number of overall events for that outcome divided by 10; predictors chosen for the final complementary log-log binary regression model for both outcomes were based on a forward selection AIC. Kaplan Meier survival curves with log rank tests were constructed to compare freedom from the composite primary outcome between groups. The optimal cut point for a continuous covariate in predicting the outcomes (survival and binary arrhythmia outcomes) was obtained as the cut point that maximizes the score statistic (equivalent to the log rank statistic for the survival outcome).^28^ A two-sided p-value of ≤0.05 was considered statistically significant. Statistical analyses were performed using SAS version 9.4 (SAS Institute, Cary, NC) and SPSS version 27 (IBM Corp, Armonk, NY).

## Results

### Baseline characteristics

The study cohort included 503 Fontan patients with median age of 15.2 (IQR 10.3, 21.3) years and 42 comparison group patients with median age of 15.7 (IQR 11.0, 19.7) years. Ventricular morphology was RV in 190 (38%), LV in 157 (31%), and mixed type in 156 (31%). The type of Fontan operations were lateral tunnel (69%), extracardiac conduit (20%), RA-PA (10%), and RA-RV (1%). Compared to the comparison group, Fontan patients had a lower BSA and were more frequently male (Table 1). Compared to the LV group, patients in the RV group were younger, had a lower BSA, and had a higher heart rate. The indications for CMR in the comparison group were family history of cardiomyopathy (n=24, 57%), concern for arrhythmogenic right ventricular dysplasia (n=3, 7%), family history of a bicuspid aortic valve (n=3, 7%), family history of sudden cardiac death (n=3, 7%), and concern for other abnormality on prior echocardiogram (n=9, 21%). The patients undergoing evaluation for cardiomyopathy had a normal CMR as well as normal clinical and genetic evaluations (when available) at most recent follow-up.

**Table 1:** Baseline characteristics, conventional CMR data, and dyssynchrony measurements for all patients and comparisons

|  | All Fontan Patients (N=503) | Comparison group LV (N=42) | Fontan Subgroups |  |  | All Fontan vs. comparison group LV | LV vs. comparison group LV |
| --- | --- | --- | --- | --- | --- | --- | --- |
|  |  |  | LV (N=157) | RV (N=190) | Mixed (N=156) |  |  |
| **Baseline characteristics** |  |  |  |  |  |  |  |
| Age at CMR, y | 15.2 (10.3-21.3) | 15.7 (11.0-19.7) | 16.6 (12.6-23.8) | 14.2 (10.0-18.8) | 14.8 (9.6-21.7) | 0.923 | 0.107 |
| Sex, male | 302 (60%) | 18 (43%) | 93 (60%) | 123 (65%) | 86 (55%) | 0.030* | 0.049* |
| BSA, m<sup>2</sup> | 1.5 (1.1-1.8) | 1.6 (1.3-1.8) | 1.6 (1.2-1.8) | 1.4 (1.0-1.7) | 1.5 (1.0-1.8) | 0.044* | 0.4436 |
| SBP, mmHg | 113(101,123) | 117 (106-124) | 114 (104-123) | 113 (102-126) | 111 (96-122) | 0.239 | 0.575 |
| SBP, z-score | 0.26 (-0.62, 1.11) | 0.50 (-0.26, 1.20) | 0.42 (-0.56, 1.09) | 0.44 (-0.37, 1.37) | 0.05 (-0.90, 0.87) | 0.287 | 0.016* |
| Heart rate, bpm | 82 (70-93) | 80 (68-89) | 78 (67-89) | 85 (73-95) | 82 (69-93) | 0.628 | 0.518 |
| %MHR | 41(36-47) | 40 (34-46) | 40 (35-45) | 43 (37-48) | 41(36-47) | 0.513 | 0.797 |
| QRS interval, ms | 100 (90-116) | 84 (78-92) | 100 (90-112) | 106 (92-120) | 100 (88-112) | <0.001* | <0.001* |
| **Conventional CMR Variables** |  |  |  |  |  |  |  |
| EDV<sub>i</sub>, mL/BSA (N=467) | 107 (92-134) | 82 (73-93) | 94 (79-115) | 115 (99-142) | 111 (95-143) | <0.001* | <0.001* |
| ESV<sub>i</sub>, mL/BSA (N=467) | 50 (39-68) | 32 (27-38) | 43 (32-54) | 56 (46-75) | 52 (39-68) | <0.001* | <0.001* |
| Mass<sub>i</sub>, grams/BSA (N=449) | 55 (46-70) | 48 (42-54) | 53 (46-66) | 56 (45-70) | 57 (47-74) | <0.001* | 0.001* |
| EF % (N=467) | 54 (47-58) | 60 (56-66) | 55 (50-60) | 51 (45-56) | 54 (46-60) | <0.001* | <0.001* |
| Indexed stroke volume (N=467) | 57 (48-66) | 51 (44-55) | 51 (44-59) | 58 (52-67) | 61 (50-72) | 0.003* | 0.502* |
| Aortic flow, L/min (N=359) | 3.0 (2.5-3.6) | 3.6 (3.2-3.8) | 3.0 (2.4-3.5) | 3.2 (2.7-3.9) | 2.9 (2.4-3.5) | 0.002* | 0.001* |
| GCS, % (N=503) | -14.6 (-16.8, | -17.9 (-19.7, | -16.6 (- | -13.6 (- | -14.0 (- | <0.001 | 0.001* |
|  | -12.2) | -17.0) | 18.5, -<br>14.3) | 15.6, -<br>11.2) | 16.3, -<br>12.0) | * |  |
| GRS, % (N=503) | 22.2 (17.66,<br>27.28) | 29.40<br>(27.29-33.63) | 26.18<br>(21.71-31.34) | 19.78<br>(15.56-24.57) | 21.20<br>(17.28-26.31) | <0.001<br>* | 0.002* |

Values are medians (interquartile range) or counts (%). LV, left ventricle; RV, right ventricle; CMR, cardiac magnetic resonance; BSA, body surface area; SBP, systolic blood pressure; %MHC, percent maximum heart rate; EDV<sub>i</sub>, indexed ventricular end-diastolic volume; ESV<sub>i</sub>, indexed ventricular end-systolic volume; Mass<sub>i</sub>, indexed ventricular mass; EF, ejection fraction; GCS, global circumferential strain; GRS, global radial strain.

### Relationship of dyssynchrony to ventricular morphology and conventional CMR metrics

Demographic, ECG, CMR, and dyssynchrony parameters for the study cohort are presented in Tables 1 and 2. In contrast to the comparison cohort, the Fontan patients had larger systemic ventricular volumes and mass, and lower EF, indexed stroke volume, GCS, GRS, and longer QRS interval. Details of all 24 CMR-derived dyssynchrony indices in the Fontan cohort and the comparison group are presented in Supplemental Table 1. Figure 2 shows the distributions for EDV_*i*_, EF, SDTTP-CS, and QRS interval across ventricular morphology types and comparison subjects. Within the Fontan cohort, the RV and mixed groups have higher volumes, lower EF, longer QRS duration, and higher SDTTP-CS. SDTTP-CS for the LV group was similar that of the comparison group LVs. Ventricular size and function measurements demonstrated modest correlation with QRS duration and SDTTP-CS (Figure 3). Among all the evaluated dyssynchrony metrics, SDTTP-CS, SDTTP-RS, and SDTTP-RSR had the highest correlation with measures of ventricular function (GCS, GRS, and EF; correlation coefficients ranging from 0.3 to 0.6; Supplemental Table 2).

**Table 2:**
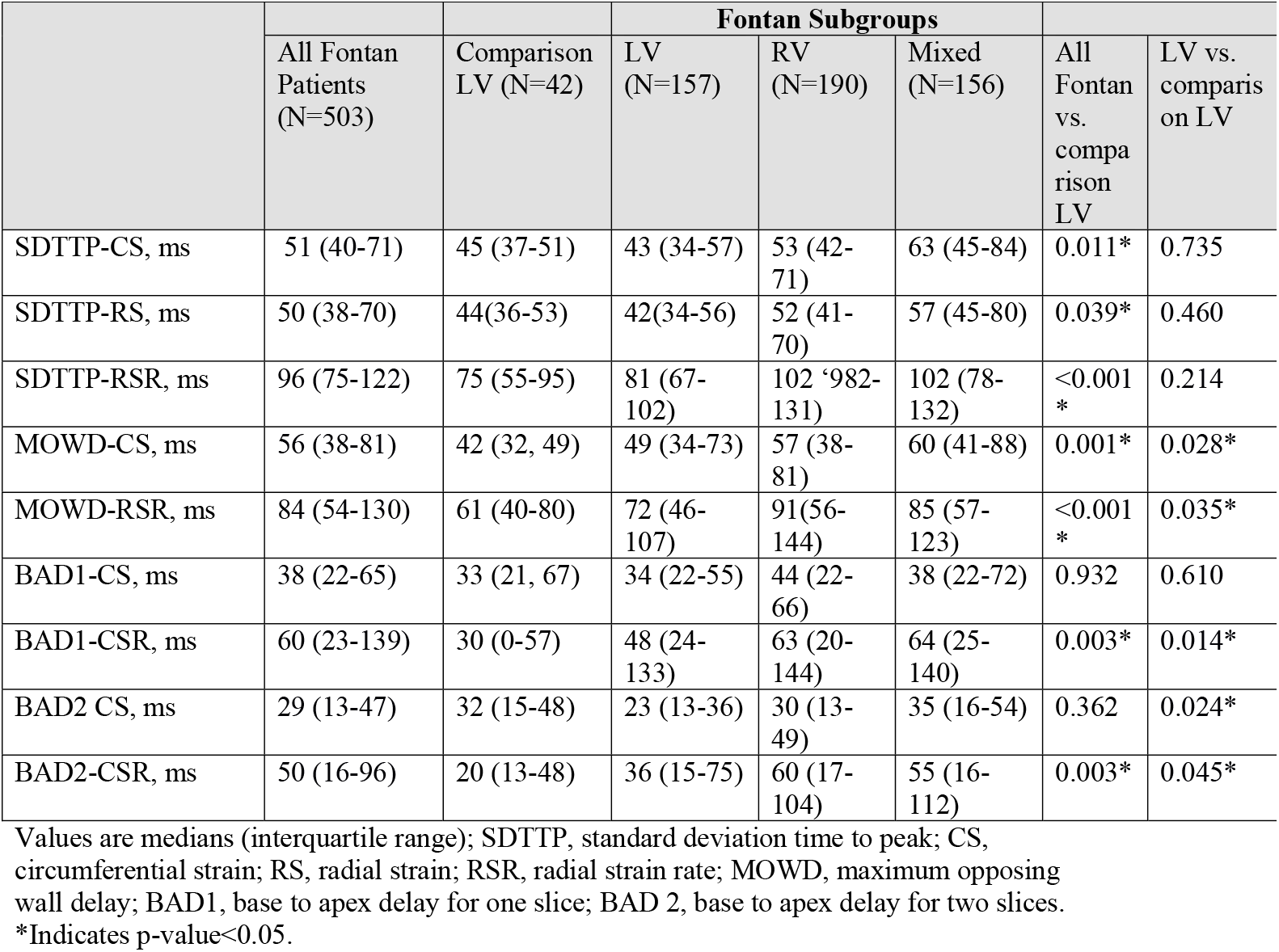
Dyssynchrony Indices

**Figure 2:**
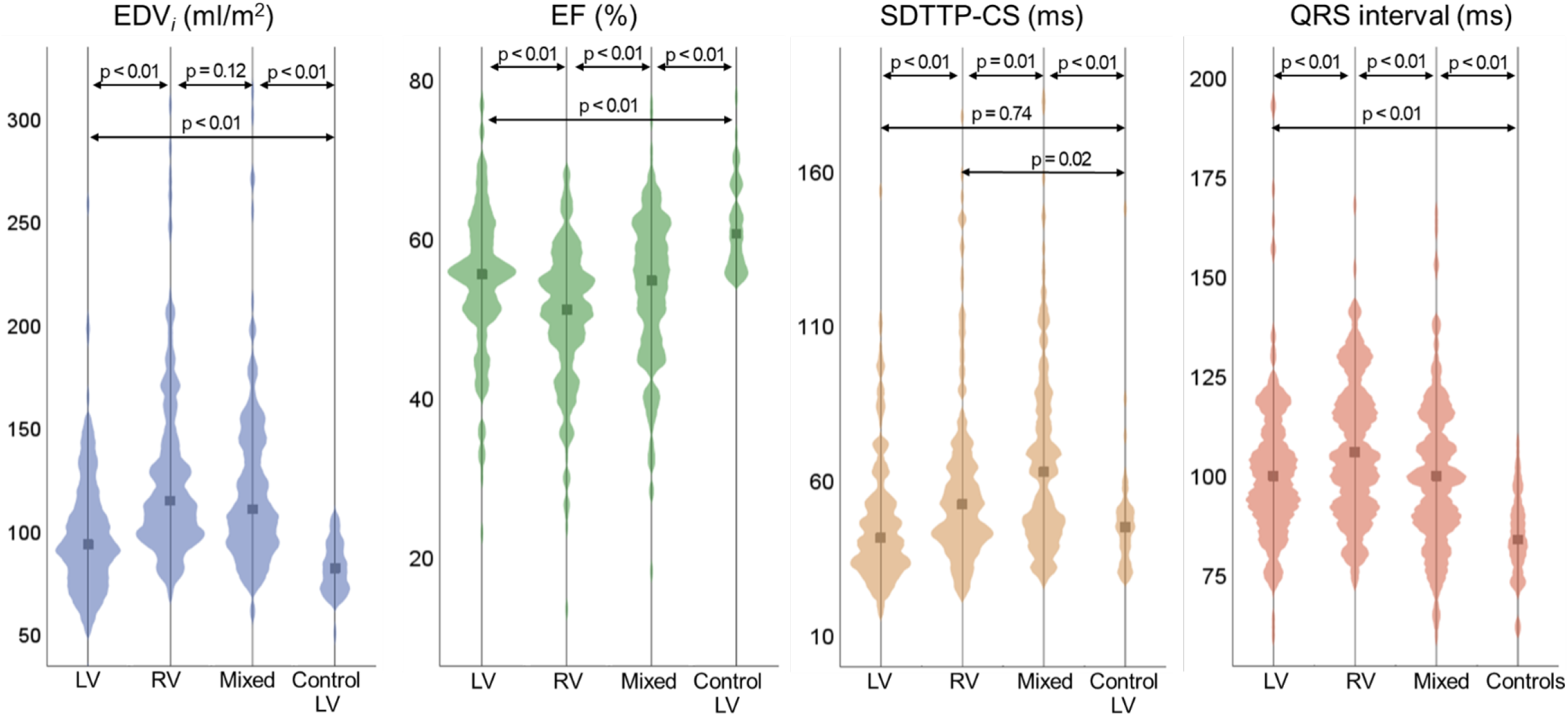
**Title:** Violin plots comparing ventricular volumes, ejection fraction, and metric of mechanical and electrical dyssynchrony by ventricular morphology **Caption:** EDV_*i*_, indexed ventricular end-diastolic volume; EF, ejection fraction, SDTTP-CS, standard deviation time to peak circumferential strain; LV, left ventricle; RV, right ventricle. *Indicates p-value<0.05.

**Figure 3:**
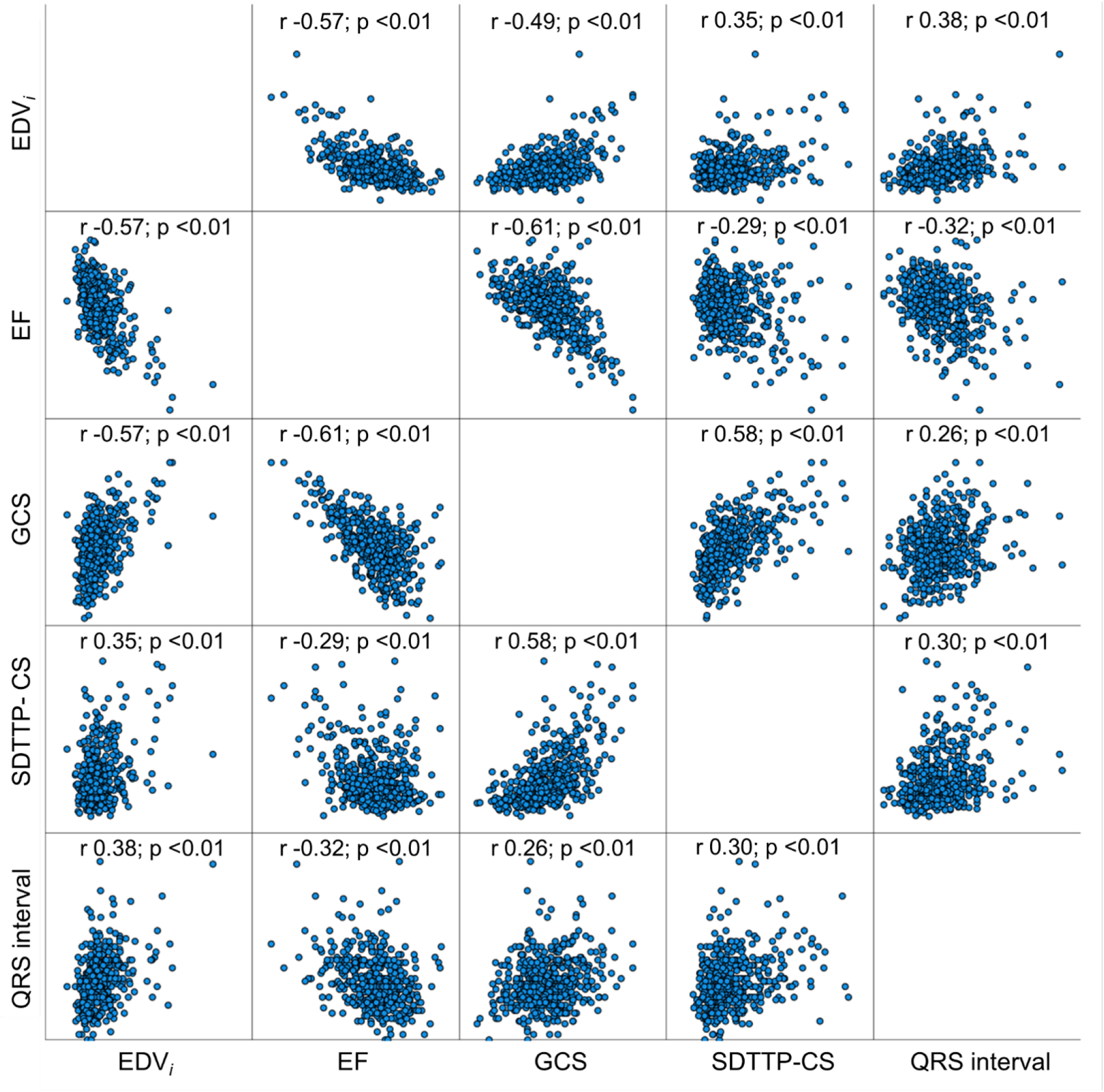
**Title:** Scatterplot matrix showing correlation between ventricular size, function, dyssynchrony, and QRS duration for the Fontan cohort (n=503) **Caption:** EDV_*i*_, indexed end diastolic volume; EF, ejection graction; GCS, global circumferential strain; SDTTP-CS, standard deviation time to peak circumferential strain

### Primary outcomes: death or heart transplantation

With a median follow-up period of 4.3 (IQR 1.3, 7.8) years, 57 (11.3%) of the patients met the composite primary outcome with 46 deaths, 8 heart transplantations, and 18 heart transplant listings. In the RV group, 28 (14.7%) patients had the outcome as compared to 19 (12.2%) in the mixed group, and 10 (6.4%) in the LV group. On bivariate Cox regression analysis, D/HTx was associated with RV or mixed ventricular morphology, lower blood pressure, higher heart rate, longer QRS, more ventricular dilation, lower EF, and higher dyssynchrony indices (Table 3). A multivariable logistic regression model found that RV morphology (HR ratio 2.3; 95% CI 1.1-4.9; p-value 0.026), EDV_*i*_ per 10 ml (HR 1.1; 95% CI 1.1-1.2; p-value <0.001) and percent maximum heart rate (MHR) [HR ratio 1.5; 95% CI 1.2-2.0; p-value 0.003] were independently associated with D/HTx, however, dyssynchrony metrics were not on multivariate analyses (Table 5). Figure 4 depicts Kaplan Meier plots for each ventricular morphology type stratified by cut off values for EDV_*i*_ and SDTTP-CS. Estimated 3-year percent D/HTx was 7.9% (95% CI 3.7-12.0) for single RVs, 3.5% (95% CI 0.4, 6.4) for mixed typed ventricles, and 1.4% (95% CI 0.0, 3.2) for single LVs (p-value = 0.005). Using the cut point of 73 ms for SDTTP-GCS that maximizes the log rank statistic, those with a SDTTP-CS of >73 ms had a higher Kaplan Meier estimate of 3-year percent D/HTx (12.5%; 95% CI 3.4, 15.5) compared to those below the cut off (3.3%; 95% CI 1.3, 5.1). Patients with EDV_*i*_ >146 ml/m^2^ had a higher rate of primary outcome than those with EDV_*i*_ <= 146 ml/m^2^ (17%; 95% CI 6.4, 23.11 vs. 2.8%; 95% CI 0.95, 4.6; p-value 0.002).

**Table 3:**
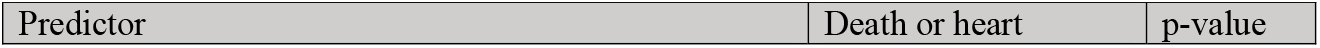
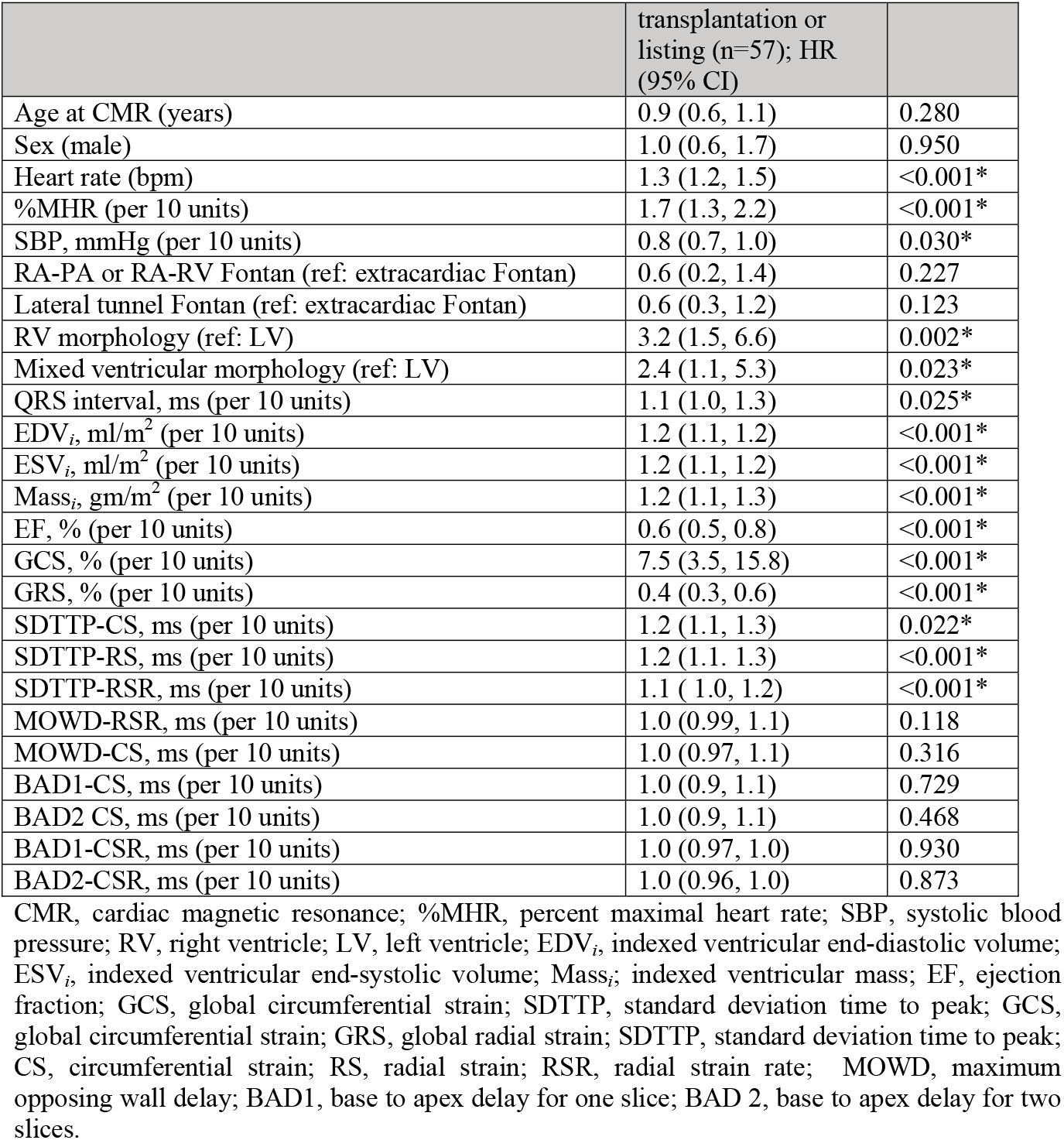
Bivariate Cox regression analysis for time to death or heart transplant listing

**Figure 4:**
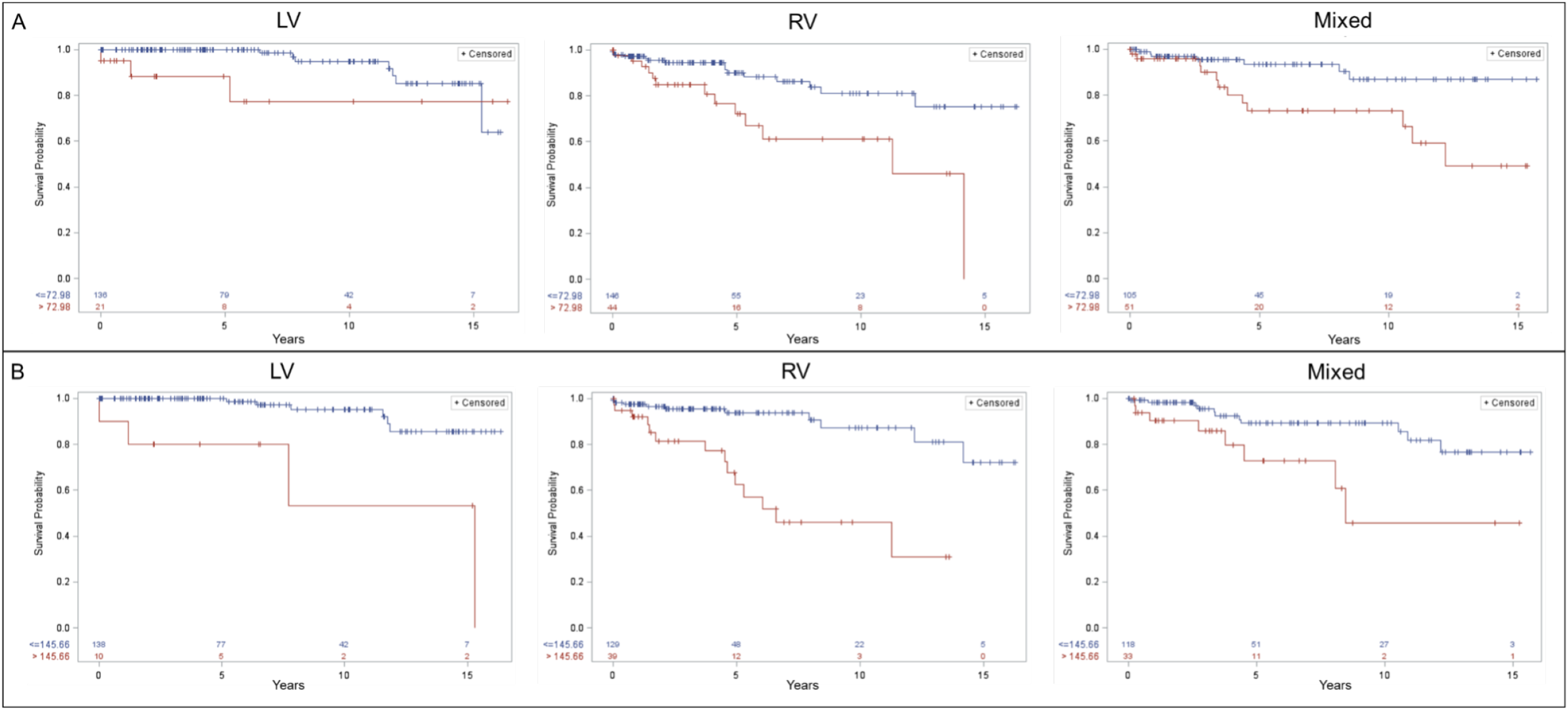
**Title:** Kaplan-Meier plots depicting freedom from the composite outcome (death, transplant, transplant listing) stratified by presence of dyssynchrony (SDTTP-CS >73 ms, panel A) and ventricular dilation (EDV_*i*_ > 146 ml/m^2^, panel B). **Caption**: Panel A: Among patients with higher dyssyynchrony, patients with LV morphology experienced the greatest freedom from the composite outcome. The overall 2df logrank p-value testing are for LV (p<0.06), RV (p<0.002) and Mixed (p=0.005). Panel B: The overall 2df logrank p-value testing are for LV (p<0.001), RV (p<0.001) and Mixed (p=0.006). The pairwise adjusted p-values using Tukey-Kramer for multiple comparisons remained significant for all tests.

### Secondary outcomes: arrhythmia rates

The total patient years follow-up used to calculate arrhythmia rates was 2807 patient-years (mean of 5.6 years per patient and median of 4.2 years per patient). The presence of significant ventricular arrhythmia (36 patients/7.2% of the cohort, giving an overall rate of 1.33 events per 100 patient-years (95% CI 0.96, 1.86) was associated with a longer QRS duration, ventricular dilation, elevated mass, lower EF, greater GCS, lower GRS, and higher dyssynchrony indices (Table 4). There was no significant difference in ventricular arrhythmias based on ventricular morphology (p-value 0.237). Patients with a QRS duration >118 ms had a rate of 2.9 ventricular arrhythmias per 100 patient-years compared to a rate of 1.0 ventricular arrhythmia per 100 patient-years for patients with QRS duration ≤118 ms (p-value = 0.001). Patients with SDTTP-CS >96 ms had a rate of 4.2 ventricular arrhythmias per 100 patient-years compared to a rate of 1.1 ventricular arrhythmias per 100 patient-years for SDTTP-CS ≤96 ms (p-value <0.001). Multivariable complementary log-log regression for rates controlling for ventricular morphology showed that EDV_*i*_ and GCS were independent predictors of ventricular arrhythmias (Table 6). Atrial arrhythmias (189 patients/37.6% of the cohort; giving an overall rate of 9.5 events per 100 patient-years (95% CI 8.2,11.1) were associated with older age, RA-PA, or RA-RV Fontan (compared to an extracardiac Fontan), longer QRS duration, greater GCS, more dilated ventricles, and elevated dyssynchrony indices (Table 4). Patients with SDTTP-CS >85 ms had a rate of 16.5 atrial arrhythmias per 100 patient-years compared to a rate of 8.6 atrial arrhythmias per 100 patient-years for SDTTP-CS ≤85 ms (p-value <0.001). Multivariable complementary log-log regression for rates showed that EDV_*i*_, GCS, and Fontan type (higher with RA-RV, RA-PA types) were independently associated with atrial arrhythmias (Table 6).

**Table 4:**
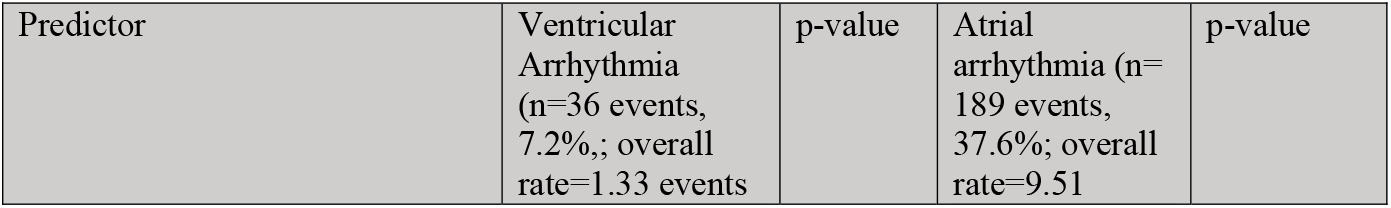
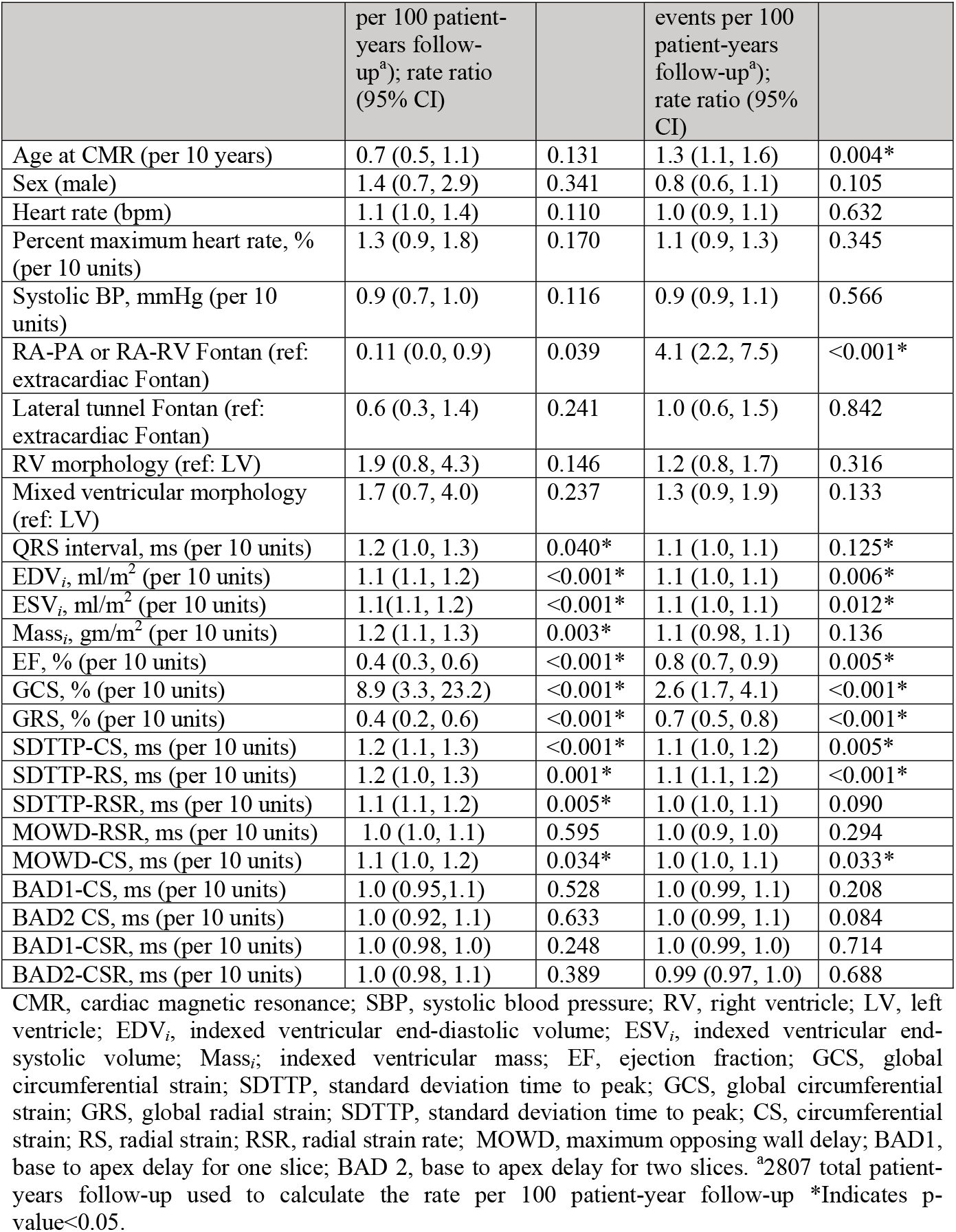
Bivariate complementary log-log binary regression models for rates a showing associations between cardiac arrhythmias and clinical and CMR variables

| Predictor | Ventricular Arrhythmia (n=36 events, 7.2%; overall rate=1.33 events | p-value | Atrial arrhythmia (n=189 events, 37.6%; overall rate=9.51 | p-value |
| --- | --- | --- | --- | --- |

|  | per 100 patient-years follow-up<sup>a</sup>; rate ratio (95% CI) |  | events per 100 patient-years follow-up<sup>a</sup>; rate ratio (95% CI) |  |
| --- | --- | --- | --- | --- |
| Age at CMR (per 10 years) | 0.7 (0.5, 1.1) | 0.131 | 1.3 (1.1, 1.6) | 0.004* |
| Sex (male) | 1.4 (0.7, 2.9) | 0.341 | 0.8 (0.6, 1.1) | 0.105 |
| Heart rate (bpm) | 1.1 (1.0, 1.4) | 0.110 | 1.0 (0.9, 1.1) | 0.632 |
| Percent maximum heart rate, % (per 10 units) | 1.3 (0.9, 1.8) | 0.170 | 1.1 (0.9, 1.3) | 0.345 |
| Systolic BP, mmHg (per 10 units) | 0.9 (0.7, 1.0) | 0.116 | 0.9 (0.9, 1.1) | 0.566 |
| RA-PA or RA-RV Fontan (ref: extracardiac Fontan) | 0.11 (0.0, 0.9) | 0.039 | 4.1 (2.2, 7.5) | <0.001* |
| Lateral tunnel Fontan (ref: extracardiac Fontan) | 0.6 (0.3, 1.4) | 0.241 | 1.0 (0.6, 1.5) | 0.842 |
| RV morphology (ref: LV) | 1.9 (0.8, 4.3) | 0.146 | 1.2 (0.8, 1.7) | 0.316 |
| Mixed ventricular morphology (ref: LV) | 1.7 (0.7, 4.0) | 0.237 | 1.3 (0.9, 1.9) | 0.133 |
| QRS interval, ms (per 10 units) | 1.2 (1.0, 1.3) | 0.040* | 1.1 (1.0, 1.1) | 0.125* |
| EDV<sub>i</sub>, ml/m<sup>2</sup> (per 10 units) | 1.1 (1.1, 1.2) | <0.001* | 1.1 (1.0, 1.1) | 0.006* |
| ESV<sub>i</sub>, ml/m<sup>2</sup> (per 10 units) | 1.1 (1.1, 1.2) | <0.001* | 1.1 (1.0, 1.1) | 0.012* |
| Mass<sub>i</sub>, gm/m<sup>2</sup> (per 10 units) | 1.2 (1.1, 1.3) | 0.003* | 1.1 (0.98, 1.1) | 0.136 |
| EF, % (per 10 units) | 0.4 (0.3, 0.6) | <0.001* | 0.8 (0.7, 0.9) | 0.005* |
| GCS, % (per 10 units) | 8.9 (3.3, 23.2) | <0.001* | 2.6 (1.7, 4.1) | <0.001* |
| GRS, % (per 10 units) | 0.4 (0.2, 0.6) | <0.001* | 0.7 (0.5, 0.8) | <0.001* |
| SDTTP-CS, ms (per 10 units) | 1.2 (1.1, 1.3) | <0.001* | 1.1 (1.0, 1.2) | 0.005* |
| SDTTP-RS, ms (per 10 units) | 1.2 (1.0, 1.3) | 0.001* | 1.1 (1.1, 1.2) | <0.001* |
| SDTTP-RSR, ms (per 10 units) | 1.1 (1.1, 1.2) | 0.005* | 1.0 (1.0, 1.1) | 0.090 |
| MOWD-RSR, ms (per 10 units) | 1.0 (1.0, 1.1) | 0.595 | 1.0 (0.9, 1.0) | 0.294 |
| MOWD-CS, ms (per 10 units) | 1.1 (1.0, 1.2) | 0.034* | 1.0 (1.0, 1.1) | 0.033* |
| BAD1-CS, ms (per 10 units) | 1.0 (0.95, 1.1) | 0.528 | 1.0 (0.99, 1.1) | 0.208 |
| BAD2 CS, ms (per 10 units) | 1.0 (0.92, 1.1) | 0.633 | 1.0 (0.99, 1.1) | 0.084 |
| BAD1-CSR, ms (per 10 units) | 1.0 (0.98, 1.0) | 0.248 | 1.0 (0.99, 1.0) | 0.714 |
| BAD2-CSR, ms (per 10 units) | 1.0 (0.98, 1.1) | 0.389 | 0.99 (0.97, 1.0) | 0.688 |

CMR, cardiac magnetic resonance; SBP, systolic blood pressure; RV, right ventricle; LV, left ventricle; EDV<sub>i</sub>, indexed ventricular end-diastolic volume; ESV<sub>i</sub>, indexed ventricular end-systolic volume; Mass<sub>i</sub>, indexed ventricular mass; EF, ejection fraction; GCS, global circumferential strain; SDTTP, standard deviation time to peak; GCS, global circumferential strain; GRS, global radial strain; SDTTP, standard deviation time to peak; CS, circumferential strain; RS, radial strain; RSR, radial strain rate; MOWD, maximum opposing wall delay; BAD1, base to apex delay for one slice; BAD 2, base to apex delay for two slices. <sup>a</sup>2807 total patient-years follow-up used to calculate the rate per 100 patient-year follow-up \*Indicates p-value<0.05.

**Table 5:** Multivariate Cox regression model for time to death or heart transplant listing. Multiple imputation used for missing data. (N=503; 57 outcomes; model c-statistic 0.79)

|  | aHR<br>(95% CI) | P-value |
| --- | --- | --- |
| RV morphology (ref: LV morphology) | 2.3 (1.1, 4.9) | 0.026* |
| Mixed ventricular morphology (ref: LV morphology) | 1.6 (0.7, 3.6) | 0.223 |
| EDV<sub>i</sub>, ml/m<sup>2</sup> (per 10 units) | 1.1 (1.1, 1.2) | <0.001* |
| Percent maximum heart rate, 10% (per 10 units) | 1.5 (1.2, 2.0) | 0.003* |

aHR, adjusted hazard ratio; RV, right ventricle; LV, left ventricle; EDV<sub>i</sub>, indexed ventricular end-diastolic volume. \*Indicates p-value<0.05.

**Table 6:** Complementary log-log binary regression models for rates (Akaike information criterion forward selection) for ventricular and atrial arrhythmias. Multiple imputation used for missing data. (N=503; 189 atrial events, 36 ventricular events; C statistic 0.702 and 0.664, respectively)

|  | **Ventricular**<br>**Arrhythmia** | **p-value** | **Atrial**<br>**Arrhythmia** | **p-value** |
| --- | --- | --- | --- | --- |
|  | aRR (95% CI) |  | aRR (95% CI) |  |
| Age at CMR (per 10 years) | 0.8 (0.5, 1.1) | 0.193 | 1.0 (0.8, 1.2) | 0.823 |
| EDV<sub>i</sub>, ml/m<sup>2</sup> (per 10 units) | 1.1 (1.05, 1.1) | <0.001* | 1.0 (1.0, 1.1) | 0.002* |
| GCS (per 10 units) | 3.4 (1.2, 10.0) | 0.023* | 1.9 (1.1, 3.2) | 0.015* |
| RA-PA or RA-RV Fontan (ref:<br>extracardiac Fontan) | -----<br>----- | ----- | 5.9 (2.8, 12.3) | <0.001* |
| Lateral tunnel Fontan (ref:<br>extracardiac Fontan) | -----<br>----- | ----- | 1.3 (0.8, 2.1) | 0.385 |

aRR, adjusted rate ratio; RA-PA, right atrium to pulmonary artery; RA-RV, right atrium to right ventricle; RV, right ventricle; LV, left ventricle; EDV<sub>i</sub>, indexed ventricular end-diastolic volume. Regression coefficients are (log of) rate ratios. \*Indicates p-value<0.05.

## Discussion

This retrospective analysis of a large cohort of patients with a Fontan circulation provides a comprehensive characterization and analysis of mechanical dyssynchrony in FSV patients using CMR-FT. The cohort covers a wide breadth of single ventricular morphology, including dominant right, left, and mixed type ventricles. Our data demonstrate a high prevalence of ventricular dyssynchrony in this population, which is associated with reduced ventricular function, cardiac arrhythmias, and D/HTx. RV and mixed morphology subtypes had a higher degree of dyssynchrony compared to those with LV morphology. Conversely, in those with LV morphology, dyssynchrony indices were more similar to those in the comparison group LVs. These results contribute to the growing body of literature demonstrating adverse ventricular remodeling in the Fontan circulation and highlight new risk factors for increased morbidity and mortality, which are critical to understanding in the management of a mounting population surviving into adulthood.

Data regarding the characterization and patterns of dyssynchrony in patients with a Fontan circulation are growing but remain to be fully elucidated. Both adult and pediatric patients with heart failure are known to have more dyssynchronous patterns of electrical and mechanical ventricular contraction, which is associated with worse LV and RV systolic function and increased morbidity.^29,30^ Similar to adults, no single gold standard measurement of mechanical dyssynchrony has gained universal acceptance or validation in pediatrics.^31,32^ Prior studies have assessed dyssynchrony in the FSV population using mid-ventricular or apical slices, typically using a 6-segment model.^19,33,34^ The majority of these are echocardiogram-based and are limited by incomplete visualization of the entire myocardium. This study builds on prior work by capturing the performance of the entire ventricle (by analyzing all segments to estimate the SDTTP), and also explores new methods of MOWD and BAD to characterize dyssynchrony. Our metrics of BAD were only modestly correlated with EF, perhaps because ventricles in the Fontan circulation have increased sphericity compared to normal bullet shaped left ventricles. While gathering normative CMR data from healthy controls remains a challenge, the inclusion of 42 age-matched comparison patients in this study sheds some light on normal dyssynchrony indices. No normative data exist for dyssynchrony measures by ventricular morphology, and there is no accepted threshold for when measurements become clinically significant. A prior study analyzed dyssynchrony in 100 patients with a Fontan circulation using long-axis views and reported that those with a SDTTP longitudinal strain of >63.5 ms was associated with the composite outcome of heart failure, unplanned hospitalization, or death.^19^ These findings are similar to the SDTTP-CS cut-off of 73 ms for D/HTx that we found in the present study.

In biventricular hearts with acquired heart diseases, QRS duration correlates with markers of ventricular dyssynchrony, dilation, and heart failure.^35,36^ QRS duration has also been identified as a prognostic indicator in CHD such as tetralogy of Fallot; although, recent studies have shown that its effect may not be independent of LV dysfunction.^37^ Similar data in the Fontan population are limited. In small echocardiography-based studies on pediatric patients with a Fontan circulation, longer QRS duration has been associated with increased incidences of dyssynchrony ^38^ with worse systolic function. Additionally, catheterization-based studies in this population have shown QRS prolongation is linked to poor hemodynamics with higher filling pressures and lower cardiac index.^39,40^ The current study analyzes a relatively large cohort and demonstrates associations between QRS duration, ventricular dyssynchrony, and adverse clinical outcomes. Our findings suggest that dyssynchrony plays an important role in the development of failing Fontan physiology and arrhythmias. Unlike previously identified unmodifiable risk factors, the contribution of CMR-based dyssynchrony metrics with poor outcomes can be informative in the management of patients with CHD. In patients with and without CHD, cardiac resynchronization therapy (CRT) has become the treatment of choice in individuals with heart failure and ventricular dyssynchrony.^41,42^ The discovery of CMR markers of mechanical dyssynchrony may offer an opportunity to identify high-risk Fontan patients who may benefit from CRT and subsequently follow their response to CRT. Furthermore, segmental analysis using FT may inform lead placement to achieve optimal synchronous contraction.

This study reaffirmed that impaired ventricular ejection fraction, ventricular dilation, and RV or mixed ventricular morphology are established risk factors for poor clinical outcomes in patients with a Fontan circulation.^3,43,44^ Multivariable analysis revealed that the association of adverse outcomes with markers of dyssynchrony is not independent of established conventional markers such as ventricular systolic dysfunction and dilation. Our findings of lower event-free survival in RV or mixed ventricular subtypes agree with prior large multicenter observational studies on Fontan patients, which have demonstrated lower long-term survival.^3^ On stratified survival analysis, however, dyssynchronous single RVs had worse outcomes compared to dyssynchronous single LVs and mixed type ventricles. This suggests that ventricular dominance remains a valuable predictor of outcomes, even amongst patients with failing Fontan physiology. Furthermore, higher heart rates were independently associated with death and heart transplantation. Perhaps the orientation of myofibers, myocardial fibrosis, and the lack of ventricular–ventricular interactions in the non-LV Fontans contribute to the development of dyssynchronous myocardial contraction, heart failure, and abnormal electrical circuits.^45–47^

Lastly, our data supported prior studies that atrial and ventricular arrhythmias are highly prevalent in patients palliated to a Fontan, with similar distribution across ventricular dominant subtypes.^7,48^ Arrhythmias were associated with ventricular dilation, dysfunction, prolonged QRS duration, Fontan type, and markers of mechanical dyssynchrony. There is lack of agreement regarding the optimal technique for the Fontan completion. The atriopulmonary Fontan has largely been abandoned as it is inefficient, results in atrial dilation, and a high incidence of atrial arrhythmias.^7^ The extracardiac Fontan has increased in popularity for its simplicity and energy efficiency, however, there are some data suggesting improved survival with the lateral tunnel Fontan.^49^ Compared to extracardiac conduit, our findings confirm the significantly increased risk with atriopulmonary Fontan with an adjusted rate ratio of 5.9 (95% CI 2.8-12.3); but also showed a relatively modestly increased risk for lateral tunnel Fontan (adjusted risk ratio 1.3, 95% CI 0.8-2.1). These findings are concordant with the published literature. In a meta-analysis of 38 publications, Zheng et al. have reported a similar early risk of atrial arrhythmias, but also an increased late risk of atrial arrhythmias with the lateral tunnel approach compared to an extracardiac conduit.^50^ These results are not surprising as the lateral tunnel approach involves multiple suture lines on the atrium and incorporates a portion of the atrium into the relatively higher-pressure Fontan baffle, either of which can act as a substrate for arrhythmia.

This study has several limitations that must be acknowledged. The generalizability of the findings to the Fontan population as a whole may be limited by single-center retrospective design that uses availability of a CMR as an inclusion criterion. The study focused on mechanical performance of the ventricles and did not analyze markers of fibrosis such as late gadolinium enhancement or myocardial T1 measurements, as that would have resulted in a substantially smaller cohort. The relatively low temporal resolution of CMR-FT analysis must also be acknowledged. Our cohort has a selection bias as pediatric patients referred for a CMR tend to be older and those with pacemakers and defibrillators were excluded. Moreover, a referral bias for CMR testing may lead to sicker and more symptomatic patients being overrepresented in the cohort. While time-to-event analysis could be conducted for the outcome of D/Htx, the same was not performed for arrhythmias. The first occurrence of an arrhythmia would be impossible to obtain accurately for the entire cohort as many patients are primarily followed at other centers and documentation in the medical records available for this study would be incomplete.

## Conclusions

Functional single ventricles in the Fontan circulation exhibit significant mechanical dyssynchrony, which is more pronounced in hearts with RV and mixed morphology compared to LV morphology. CMR-derived dyssynchrony indices correlate with ventricular size and function and are associated with death or need for heart transplantation as well as cardiac arrhythmias. These data add to the growing understanding of progressive decline in ventricular performance in the Fontan population.

## Data Availability

Data is available upon request

## Abbreviations

AIC: Akaike information criterion
BAD: base-to-apex delay
CMR: cardiac magnetic resonance
CO_*i*_: indexed cardiac output
CS: circumferential strain
D/HTx: death or heart transplant listing
ECG: electrocardiogram
EDV: end-diastolic volume
EDV_*i*_: indexed end-diastolic volume
EF: ejection fraction
ESV_*i*_: indexed end-systolic volume
FSV: functional single ventricle
FT: feature tracking
GCD: global circumferential displacement
GCS: global circumferential strain
GCSR: global circumferential strain rate
GRD: global radial displacement
GRS: global radial strain
GRSR: global radial strain rate
IQR: interquartile ranges
LV: left ventricle
Mass_i_: indexed ventricular mass
MHR: maximum heart rate
MOWD: maximum opposing wall delay
RS: radial strain
RV: right ventricle
SDTTP: standard deviation of time-to-peak
SV_*i*_: indexed stroke volume
VT: ventricular tachycardia

## Acknowledgments

The authors would like to acknowledge Kai-Ou Tang for her assistance with the central illustration.

## References

1. Diller GP, Kempny A, Alonso-Gonzalez R, Swan L, Uebing A, Li W, et al. Survival Prospects and Circumstances of Death in Contemporary Adult Congenital Heart Disease Patients under Follow-Up at a Large Tertiary Centre. Circulation 2015. doi:10.1161/CIRCULATIONAHA.115.017202.

2. Idorn L, Olsen M, Jensen AS, Juul K, Reimers JI, Sørensen K, et al. Univentricular hearts in Denmark 1977 to 2009: Incidence and survival. Int J Cardiol 2013. doi:10.1016/j.ijcard.2012.03.182.

3. Oster ME, Knight JH, Suthar D, Amin O, Kochilas LK. Long-term outcomes in single-ventricle congenital heart disease importance of ventricular morphology. Circulation 2018. doi:10.1161/CIRCULATIONAHA.118.036821.

4. Pundi KN, Johnson JN, Dearani JA, Pundi KN, Li Z, Hinck CA, et al. 40-Year Follow-Up After the Fontan Operation. J Am Coll Cardiol 2015. doi:10.1016/j.jacc.2015.07.065.

5. Stephenson EA, Lu M, Berul CI, Etheridge SP, Idriss SF, Margossian R, et al. Arrhythmias in a Contemporary Fontan Cohort: Prevalence and Clinical Associations in a Multi-Center Cross-Sectional Study. J Am Coll Cardiol 2010;56:890–896. doi:10.1016/j.jacc.2010.03.079.

6. Alsaied T, Bokma JP, Engel ME, Kuijpers JM, Hanke SP, Zuhlke L, et al. Factors associated with long-term mortality after Fontan procedures: a systematic review. Heart 2017;103:104– 110. doi:10.1136/heartjnl-2016-310108.

7. Khairy P, Fernandes SM, Mayer JE, Triedman JK, Walsh EP, Lock JE, et al. Long-term survival, modes of death, and predictors of mortality in patients with Fontan surgery. Circulation 2008. doi:10.1161/CIRCULATIONAHA.107.738559.

8. Giannakoulas G, Dimopoulos K, Yuksel S, Inuzuka R, Pijuan-Domenech A, Hussain W, et al. Atrial tachyarrhythmias late after Fontan operation are related to increase in mortality and hospitalization. Int J Cardiol 2012. doi:10.1016/j.ijcard.2010.12.049.

9. Zafrir N, Nevzorov R, Bental T, Strasberg B, Gutstein A, Mats I, et al. Prognostic value of left ventricular dyssynchrony by myocardial perfusion-gated SPECT in patients with normal and abnormal left ventricular functions. J Nucl Cardiol 2014. doi:10.1007/s12350-014-9852-1.

10. Gowda ST, Ahmad A, Younoszai A, Du W, Singh HR, Pettersen MD, et al. Left ventricular systolic dyssynchrony in pediatric and adolescent patients with congestive heart failure. J Am Soc Echocardiogr 2012. doi:10.1016/j.echo.2012.01.007.

11. Al-Biltagi MA, Abd Rab Elrasoul Tolba O, El Mahdy H, Donia A, Elbanna S. Echocardiographic assessment of left ventricular dyssynchrony in Egyptian children with congestive heart failure due to dilated cardiomyopathy. Cardiol Young 2015. doi:10.1017/S1047951114001863.

12. Aljaroudi W, Aggarwal H, Venkataraman R, Heo J, Iskandrian AE, Hage FG. Impact of left ventricular dyssynchrony by phase analysis on cardiovascular outcomes in patients with end-stage renal disease. J Nucl Cardiol 2010. doi:10.1007/s12350-010-9271-x.

13. Jiang W, Liu Y, He Z, Zhou Y, Wang C, Jiang Z, et al. Prognostic value of left ventricular mechanical dyssynchrony in hypertrophic cardiomyopathy patients with low risk of sudden cardiac death. Nucl Med Commun 2021. doi:10.1097/MNM.0000000000001322.

14. Modin D, Biering-Sørensen SR, Møgelvang R, Jensen JS, Biering-Sørensen T. Prognostic Importance of Left Ventricular Mechanical Dyssynchrony in Predicting Cardiovascular Death in the General Population. Circ Cardiovasc Imaging 2018;11:e007528. doi:10.1161/CIRCIMAGING.117.007528.

15. Onishi T, Saha SK, Ludwig DR, Onishi T, Marek JJ, Cavalcante JL, et al. Feature tracking measurement of dyssynchrony from cardiovascular magnetic resonance cine acquisitions: Comparison with echocardiographic speckle tracking. J Cardiovasc Magn Reson 2013. doi:10.1186/1532-429X-15-95.

16. Friedberg MK, Roche SL, Mohammed AF, Balasingam M, Atenafu EG, Kantor PF. Left ventricular diastolic mechanical dyssynchrony and associated clinical outcomes in children with dilated cardiomyopathy. Circ Cardiovasc Imaging 2008. doi:10.1161/CIRCIMAGING.108.782086.

17. Friedberg MK, Silverman NH, Dubin AM, Rosenthal DN. Right Ventricular Mechanical Dyssynchrony in Children with Hypoplastic Left Heart Syndrome. J Am Soc Echocardiogr 2007. doi:10.1016/j.echo.2007.02.015.

18. Friedberg MK, Roche SL, Mohammed AF, Balasingam M, Atenafu EG, Kantor PF. Left ventricular diastolic mechanical dyssynchrony and associated clinical outcomes in children with dilated cardiomyopathy. Circ Cardiovasc Imaging 2008;1:50–57. doi:10.1161/CIRCIMAGING.108.782086.

19. Ishizaki U, Nagao M, Shiina Y, Inai K, Mori H, Takahashi T, et al. Global strain and dyssynchrony of the single ventricle predict adverse cardiac events after the Fontan procedure: Analysis using feature-tracking cine magnetic resonance imaging. J Cardiol 2019. doi:10.1016/j.jjcc.2018.07.005.

20. Samyn MM, Powell AJ, Garg R, Sena L, Geva T. Range of ventricular dimensions and function by steady-state free precession cine MRI in repaired tetralogy of fallot: Right ventricular outflow tract patch vs. conduit repair. J Magn Reson Imaging 2007. doi:10.1002/jmri.21094.

21. Harrington JK, Ghelani S, Thatte N, Valente AM, Geva T, Graf JA, et al. Impact of pulmonary valve replacement on left ventricular rotational mechanics in repaired tetralogy of Fallot. J Cardiovasc Magn Reson Off J Soc Cardiovasc Magn Reson 2021;23:61. doi:10.1186/s12968-021-00750-3.

22. Tanaka H, Monahan KD, Seals DR. Age-predicted maximal heart rate revisited. J Am Coll Cardiol 2001;37:153–156. doi:10.1016/s0735-1097(00)01054-8.

23. Lipsitz SR, Fitzmaurice GM, Weiss RD. Using Multiple Imputation with GEE with Non-monotone Missing Longitudinal Binary Outcomes. Psychometrika 2020. doi:10.1007/s11336-020-09729-y.

24. Horton NJ, Lipsitz SR. Multiple imputation in practice: Comparison of software packages for regression models with missing variables. Am Stat 2001. doi:10.1198/000313001317098266.

25. Draper CC, Voller A, Carpenter RG. The epidemiologic interpretation of serologic data in malaria. Am J Trop Med Hyg 1972;21:696–703. doi:10.4269/ajtmh.1972.21.696.

26. Martuzzi M, Elliott P. Estimating the incidence rate ratio in cross-sectional studies using a simple alternative to logistic regression. Ann Epidemiol 1998;8:52–55. doi:10.1016/s1047-2797(97)00106-3.

27. Piegorsch WW. Complementary Log Regression for Generalized Linear Models. Am Stat 1992;46:94–99. doi:10.1080/00031305.1992.10475858.

28. Contal C, O’Quigley J. An application of changepoint methods in studying the effect of age on survival in breast cancer. Comput Stat Data Anal 1999. doi:10.1016/S0167-9473(98)00096-6.

29. Bleeker GB, Bax JJ, Steendijk P, Schalij MJ, van der Wall EE. Left ventricular dyssynchrony in patients with heart failure: Pathophysiology, diagnosis and treatment. Nat Clin Pract Cardiovasc Med 2006. doi:10.1038/ncpcardio0505.

30. Tanaka H, Nesser HJ, Buck T, Oyenuga O, Jánosi RA, Winter S, et al. Dyssynchrony by speckle-tracking echocardiography and response to cardiac resynchronization therapy: Results of the Speckle Tracking and Resynchronization (STAR) study. Eur Heart J 2010. doi:10.1093/eurheartj/ehq213.

31. Chung ES, Leon AR, Tavazzi L, Sun JP, Nihoyannopoulos P, Merlino J, et al. Results of the predictors of response to crt (prospect) trial. Circulation 2008. doi:10.1161/CIRCULATIONAHA.107.743120.

32. Faletra FF, Conca C, Klersy C, Klimusina J, Regoli F, Mantovani A, et al. Comparison of Eight Echocardiographic Methods for Determining the Prevalence of Mechanical Dyssynchrony and Site of Latest Mechanical Contraction in Patients Scheduled for Cardiac Resynchronization Therapy. Am J Cardiol 2009. doi:10.1016/j.amjcard.2009.02.043.

33. Rösner A, Khalapyan T, Dalen H, McElhinney DB, Friedberg MK, Lui GK. Classic-Pattern Dyssynchrony in Adolescents and Adults With a Fontan Circulation. J Am Soc Echocardiogr Off Publ Am Soc Echocardiogr 2018;31:211–219. doi:10.1016/j.echo.2017.10.018.

34. Goudar S, Forsha D, White DA, Sherman A, Shirali G. Single ventricular strain measures correlate with peak oxygen consumption in children and adolescents with Fontan circulation. Cardiol Young 2022:1–7. doi:10.1017/S1047951122002323.

35. Shaik SA, Oruganti SS. Relationship of echocardiographic left ventricular dyssynchrony with QRS width on surface electrocardiogram in patients with systolic heart failure: An observational study. Indian Heart J 2021. doi:10.1016/j.ihj.2021.08.004.

36. Biering-Sørensen T, Shah SJ, Anand I, Sweitzer N, Claggett B, Liu L, et al. Prognostic importance of left ventricular mechanical dyssynchrony in heart failure with preserved ejection fraction. Eur J Heart Fail 2017. doi:10.1002/ejhf.789.

37. Krieger EV, Zeppenfeld K, DeWitt ES, Duarte VE, Egbe AC, Haeffele C, et al. Arrhythmias in Repaired Tetralogy of Fallot: A Scientific Statement From the American Heart Association. Circ Arrhythm Electrophysiol 2022;15:e000084. doi:10.1161/HAE.0000000000000084.

38. Zhong S, Zhang Y-Q, Chen L, Zhang Z-F, Wu L-P, Hong W. Ventricular function and dyssynchrony in children with a functional single right ventricle using real time three-dimensional echocardiography after fontan operation. Echocardiography 2021;38:1218–1227. doi:10.1111/echo.15148.

39. Gokhale J, Husain N, Nicholson L, Texter KM, Zaidi AN, Cua CL. QRS duration and mechanical dyssynchrony correlations with right ventricular function after fontan procedure. J Am Soc Echocardiogr 2013. doi:10.1016/j.echo.2012.10.018.

40. Wilson LH, Chowdhury SM, Jackson LB. QRS fragmentation and prolongation as predictors of exercise capacity in patients after Fontan palliation. Pacing Clin Electrophysiol PACE 2022;45:786–796. doi:10.1111/pace.14514.

41. Dubin AM, Janousek J, Rhee E, Strieper MJ, Cecchin F, Law IH, et al. Resynchronization therapy in pediatric and congenital heart disease patients: An international multicenter study. J Am Coll Cardiol 2005. doi:10.1016/j.jacc.2005.05.096.

42. Chubb H, Bulic A, Mah D, Moore JP, Janousek J, Fumanelli J, et al. Impact and Modifiers of Ventricular Pacing in Patients With Single Ventricle Circulation. J Am Coll Cardiol 2022;80:902–914. doi:10.1016/j.jacc.2022.05.053.

43. Ghelani SJ, Colan SD, Azcue N, Keenan EM, Harrild DM, Powell AJ, et al. Impact of Ventricular Morphology on Fiber Stress and Strain in Fontan Patients. Circ Cardiovasc Imaging 2018;11. doi:10.1161/CIRCIMAGING.117.006738.

44. Ghelani SJ, Lu M, Sleeper LA, Prakash A, Castellanos DA, Clair NS, Powell AJ Rrh. Longitudinal changes in ventricular size and function are associated with death and transplantation late after the Fontan operation. J Cardiovasc Magn Reson 2022. doi:10.1186/s12968-022-00884-y.

45. Fogel MA, Weinberg PM, Gupta KB, Rychik J, Hubbard A, Hoffman EA, et al. Mechanics of the single left ventricle: a study in ventricular-ventricular interaction II. Circulation 1998;98:330–338. doi:10.1161/01.cir.98.4.330.

46. Tham EB, Smallhorn JF, Kaneko S, Valiani S, Myers KA, Colen TM, et al. Insights into the Evolution of Myocardial Dysfunction in the Functionally Single Right Ventricle between Staged Palliations Using Speckle-Tracking Echocardiography. J Am Soc Echocardiogr 2014;27:314–322. doi:10.1016/j.echo.2013.11.012.

47. Fogel MA, Gupta KB, Weinberg PM, Hoffman EA. Regional wall motion and strain analysis across stages of Fontan reconstruction by magnetic resonance tagging. Am J Physiol - Heart Circ Physiol 1995;269:H1132–H1152.

48. Çelik Dr. M, Saritaş B, Tatar T, Özkan M, Akay T, Aşlamaci S. Risk factors for postoperative arrhythmia in patients with physiologic univentricular hearts undergoing Fontan procedure. Anadolu Kardiyol Derg 2012. doi:10.5152/akd.2012.099.

49. Weixler VHM, Zurakowski D, Kheir J, Guariento A, Kaza AK, Baird CW, et al. Fontan with lateral tunnel is associated with improved survival compared with extracardiac conduit. J Thorac Cardiovasc Surg 2020;159:1480-1491.e2. doi:10.1016/j.jtcvs.2019.11.048.

50. Zheng J, Li Z, Li Q, Li X. Meta-analysis of Fontan procedure. Herz 2018;43:238–245. doi:10.1007/s00059-017-4553-6.

